# The Association between Women’s Perception of Community Support for and Utilization of Maternity Healthcare Services in Ethiopia

**DOI:** 10.1101/2025.09.25.25336698

**Authors:** Yongyi Lu, Sally Safi, Solomon Shiferaw, Linnea A. Zimmerman

## Abstract

Ethiopia has one of the highest maternal mortality ratios in sub-Saharan Africa. Many factors contribute, including limited access to and use of maternity care services. Community support plays an important role in influencing women’s utilization of such services. The objective of this study was to analyze the association between women’s perception of community support and their utilization of maternity healthcare services in Ethiopia while exploring how this association varies by urban and rural residence. Longitudinal data from the Performance Monitoring in Action Ethiopia was used. We excluded women who were postpartum at baseline, did not complete the six-week follow-up survey, and did not deliver a live birth. The total analytic sample for this study was 1,924. We used logistic regression to analyze the relationship between a woman’s perception of community support for the relevant component and the service utilization. Then, we included an interaction term between community perceptions and residence for each model. The proportion of women with four or more antenatal care visits, who gave birth in a health facility, and had postnatal care visits within 2 days postpartum are 49%, 52%, and 41%, respectively. Women’s access to comprehensive maternity care was 25%. Women who perceived their communities as “fully supportive” of comprehensive maternity care were about twice as likely to receive such care compared to women who perceived that the community was not fully supportive of comprehensive maternity care (aOR: 1.89, 95% CI: 1.49-2.38). Regarding the full continuum of care, urban women who perceive full support were significantly more likely to receive all components of care. Perceived community support is an important predictor of women’s utilization of maternal care in Ethiopia. These findings highlight a key factor influencing care-seeking behavior and variation between urban and rural residence, contributing to ongoing disparities in healthcare access.

## Introduction

Sub-Saharan Africa has the highest burden of maternal mortality in the world, with around 85% of maternal deaths (1). Ethiopia, specifically, has one of the highest maternal mortality ratios in sub-Saharan Africa, with approximately 401 maternal deaths per 100,000 live births, recording the fourth largest number of maternal deaths in the world in 2020 (2–4). Many factors contribute to maternal mortality in Ethiopia, such as lack of transportation to health facilities, shortage of life-saving maternal supplies in health facilities, and lack of access to and utilization of the continuum of maternity care services, which includes four or more antenatal care (ANC) visits, use of facility-based delivery services, and receipt of postnatal care (PNC) for the mother within 48 hours of delivery (5–7).

ANC is widely considered an essential service during pregnancy (8). ANC includes prevention, such as iron supplementation, toxoid immunization; treatment, such as treatment for sexually transmitted infections; and health education (9,10). Currently, Ethiopia has undertaken several intervention programs to promote ANC access and improve the quality of ANC services such as conducting health extension programs, having an organized community health structure, and implementing community-based health insurance (11). Recent estimates suggest that despite these efforts, ANC utilization remains low; estimates from the most recent DHS suggest that approximately 74% of women received at least one ANC visit from a skilled provider and only 43% received four or more visits (12). These figures also mask significant geographic variability, with approximately 84% of urban women receiving at least one visit and 59% receiving four or more relative to 71% and 37%, respectively, of rural women (12).

Delivering in a health facility, with a skilled birth attendant, is the second critical component of the continuum of care. Evidence suggests that prioritizing intrapartum care, that is ensuring that women deliver with a skilled attendant in a facility that is ready to address an obstetric emergency, can significantly reduce maternal morbidity and mortality (13). Given the effectiveness of facility delivery, the WHO has long recommended that pregnant women give birth in a health facility to prevent maternal mortality (14). In Ethiopia, the Federal Ministry of Health introduced a policy of free delivery services in public health facilities in 2013 (15). However, the use of healthcare facilities for delivery is still very low in Ethiopia. According to the 2019 Ethiopia Demographic and Health Survey (DHS), only 48% of women who delivered a live birth in the five years prior to the survey were delivered in a health facility (12). As with ANC utilization, these numbers mask significant variation with almost 80% of urban women delivering in a facility relative to 20% of rural women.

Globally, approximately 57% of maternal deaths occur in the postpartum period within 6 weeks (7). In Ethiopia, around 51% to 75% of maternal deaths occur during the postpartum period (7). To reduce maternal and infant deaths, the WHO suggests that both mother and infant should receive a PNC visit within one day (24 hours) of birth, no matter where the baby is born, and have a minimum of four PNC visits within the first six weeks after birth (16). Visits should include screening for obstetric complications, prevention of infectious diseases, general well-being management, breastfeeding promotion, and discussion of postpartum family planning options (17). PNC visits are also important opportunities to check mother and baby for danger signs such as low body temperature, shortness of breath, and fever (18). According to the 2019 Ethiopia DHS, only 35% of newborns and 34% of women received PNC services within two days of birth, again with significant geographic disparity (48% of urban women received a PNC check for their health versus 29% of rural women) (12). There is evidence that utilization of PNC services is increasing, however, overall use remains low; the utilization of PNC services among women increased from 17% in 2016 to 34% in 2019 (19).

Due to the generally low utilization of each component service, utilization of the entire continuum of maternal health services – four or more ANC visits, facility delivery, and receipt of a PNC visit within 48 hours after birth – remains well below optimal levels. Estimates in Ethiopia range from 9% to 42 %, with urban women having approximately 2.8 times higher odds of completing the continuum than their rural counterparts (20).

A number of barriers have been identified that impact use of each component service, in addition to the entire continuum of care. ANC utilization is negatively impacted by both supply-side factors, such as shortage of required materials and disrespectful care by providers, and demand-side factors, such as transport costs and partner approval (21). Reasons for the low rate of use of health facilities include a perceived high cost of delivery, lack of transportation, shortage of supplies, low level of awareness about complications of pregnancy, and cultural barriers (15). Some barriers to accessing PNC include lack of funding for PNC, lack of PNC priorities from obstetricians, and lack of supplies for PNC provision (22).

Qualitative evidence suggests that community and cultural factors also impact receipt of maternal care services (21,23). One study in the Amhara region of Ethiopia found that communities supported facility delivery through taking care of children, managing household chores, and supporting transport costs and logistics (24). The study did not, however, explore whether or how normative support for different maternal health services affected use. Another study in areas surrounding Addis Ababa, the capital, found that community views of quality and utility of antenatal services were impactful on both women’s initial and return attendance (21). Community norms that facility delivery should only be used in case of emergency have been found to support high rates of home delivery in pastoralist communities (25). Conversely, as utilization of services has increased, more women may have had positive experiences, leading to higher community support for services (26).

This qualitative evidence identifies ways that community factors can both inhibit and support utilization of maternity care services, but few quantitative studies in Ethiopia have explored this question, and none in relation to the effect of community support across the entire continuum of care. Teferi and colleagues found that if a woman perceived more community support for facility delivery, she had higher odds of expressing a preference for facility delivery, but they did not assess whether perceived community support was associated with facility delivery itself (27). Mamo and colleagues found that women who perceived greater support from partners, family members, and friends to deliver in a facility were significantly more likely to do so (28). Significant gaps remain, however, as few studies have explored the effect of community support on postnatal care, on the continuum of care more broadly, or how the influence of community support may differ by residence and contribute to the differentials in utilization by urban and rural residence identified above.

Due to persistent maternal mortality and poor utilization of ANC, facility-based delivery, and PNC in Ethiopia, many previous studies have analyzed other factors affecting the utilization of these maternity healthcare services such as education, mass media, and family size (29,30). However, to our knowledge, no study has examined how women’s perception of community support influences the utilization of each component and the entirety of the continuum of care in Ethiopia, nor whether these differ by residence. It is important to understand community-level support of ANC, PNC, and facility-based delivery and how this may affect women’s utilization of these healthcare services. Our objective is thus to 1) identify the relationship between perceived community support for each service (ANC, facility delivery, and PNC) with receipt of the service; 2) to identify whether greater community support is associated with completing the entire continuum of care; and 3) whether the influence of each differs by urban versus rural residence.

## Materials and Methods

### Ethical Clearance

All procedures were reviewed and approved by Addis Ababa University, College of Health Sciences (AAU/CHS) (Ref: AAUMF 01-008) and the Johns Hopkins University Bloomberg School of Public Health (JHSPH) Institutional Review Board (FWA00000287) and carried out in accordance with relevant guidelines and regulations. Verbal informed consent was obtained from all of the participants. The IRB approves verbal consent procedures (without a need for written consent) for simple surveys without any invasive procedures in an environment where literacy is low. Women under the age of 18 who are married are considered emancipated minors and are able to provide informed consent. No unemancipated minors were included in this survey.

### Data sources

This longitudinal study used data from the Performance Monitoring for Action (PMA) Ethiopia project. PMA Ethiopia is a collaboration between the Johns Hopkins Bloomberg School of Public Health, the Ethiopian Federal Ministry of Health (FMoH), and Addis Ababa University (AAU). PMA Ethiopia implemented a longitudinal survey of pregnant and postpartum women, which focused on maternal and newborn health and reproductive health topics (31). Women were enrolled during pregnancy or within six weeks postpartum and completed baseline interviews at enrollment and follow-up interviews at six weeks, six months and one year postpartum.

Two datasets from the PMA Ethiopia panel survey were used for this analysis: the baseline survey, conducted from September 15, 2019 to December 30, 2019, and the 6-week follow-up survey, conducted throughout September 22, 2019 to June 26, 2020. The panel study was conducted using a multi-stage design in six regions - Addis Ababa, Afar, Amhara, Oromia, Tigray, and Southern Nations, Nationalities, and People’s region.^1^ Urban and rural stratification was applied within Amhara, Oromia, Tigray and SNNP. In the first stage of sampling, a total of 216 enumeration areas (EAs), were randomly selected using probability-proportional-to-size. In the second stage of sampling, all households in the selected EAs were listed and census of all household members was conducted. All women between the ages of 15-49 were screened for eligibility (self-reported pregnancy or less than six weeks postpartum) and, if eligible, were invited to participate in the survey. At the baseline interview, resident enumerators explained the study’s purpose and read the approved consent language to the women approached for consent (31). In accordance with the National Research Ethics Review Guidelines in Ethiopia, oral consent was granted due to widespread illiteracy (32). During each follow-up, women were asked if they still agreed to participate and if they had any questions (31). Women under age 18 are considered to be emancipated minors within Ethiopia if they are married. No unemancipated minors were included in this study.

A total of 2,855 women participated in the baseline interview and 2,669 women participated in the 6-week follow-up survey. The exclusion criteria in this study were women who were postpartum at baseline (n=616), did not complete the six-week follow-up survey (n=175), and who did not deliver a live birth (n=140). The total analytic sample for this study was 1,924.

### Measures Outcome measures

Our dependent variables (outcomes) were based on survey questions that assessed women’s utilization of maternity care. Using the 6-week follow-up survey, we assessed four measures:

1. receipt of four or more ANC visits from a trained provider, including a health extension worker (HEW),
2. delivered in a health facility, and
3. received PNC within 2 days of giving birth. Women were classified as receiving PNC if a trained provider visited them in their home and discussed their health, they visited a trained provider at a facility and discussed their health, or in the case of women who delivered in a facility, if someone checked on their health prior to discharge.
4. receipt of all three of the above components comprehensive maternity care.

### Key independent variables

Our independent variables (predictors) were based on categorical measures from the baseline survey assessing women’s perception of community support for specific maternity healthcare services. Specifically, we assessed three measures using the following questions from the baseline survey:

(1) Do most, some, few, or no people in your community encourage women to **<u>deliver at</u>**

**a facility?**

(2) Do most, some, few, or no people in your community encourage going to **<u>antenatal</u>**

**care?**

(3) Do most, some, few, or no people in your community encourage women to seek **<u>postnatal care</u>**?

The response options for the above three questions included: no people, few people, some people, and most people. For our analysis of each specific service, we collapsed the first two categories into “no or few people.” “Do Not Know” was included as a potential option, however, the number of “do not know” responses ranged from 1.46% (n=28) for community support of antenatal care to 2.08% (n=40) for community support of postnatal care. The small sample sizes did not allow for sufficient power to detect differences in this group and thus, were treated as missing.

We also created a dichotomous measure for perceived community support for all types of maternity care. Respondents who answered “most people” for each of the above three questions were classified into a “High support” category and all other respondents were classified in a “Lower support” category.

### Adjustment variables

We identified sociodemographic variables that we hypothesized were likely to confound the relationship of perceived community support and receipt of each service, including maternal age (categorical variable in five-year age groups), education (categorical variable indicating none, primary, secondary and above), household wealth quintile, and region. Due to almost universal marriage in this population of currently pregnant women, we did not adjust by marital status.

### Analysis

Exploratory analyses were used to describe the study sample and summarize missingness. For each component of the continuum of care and for the entire continuum, we used logistic regression to analyze the relationship between a woman’s perception of community support for the relevant component and the utilization of that service. To determine if the influence of community support differed by urban versus rural residence, we then included an interaction term between community perceptions and residence for each model. All analyses accounted for complex survey design through the application of survey weights and accounting for clustering within EAs. All analyses were conducted using Stata 18 SE (33).

## Results

**Table 1** shows the distribution of characteristics of the sample, with weighted percentages. About half of the women (55%) were aged 20-29 and 42% had never attended school. In terms of perceptions of community support for maternity care, forty-nine percent (49%) of respondents indicated that they felt that most people in their community encouraged pregnant women to seek ANC. Fifty-two percent (52%) of respondents felt that most people in their community encouraged pregnant women to give birth in a health facility. Forty-one percent (41%) felt that most people in the community encouraged women who had given birth to seek PNC. In terms of use of maternity care, 44% of respondents indicated that they had 4 or more ANC visits, 55% indicated that they delivered in a health facility, 39% reported they had a PNC visit within 2 days and 25% reported they had received all these services.

**Table 1.**
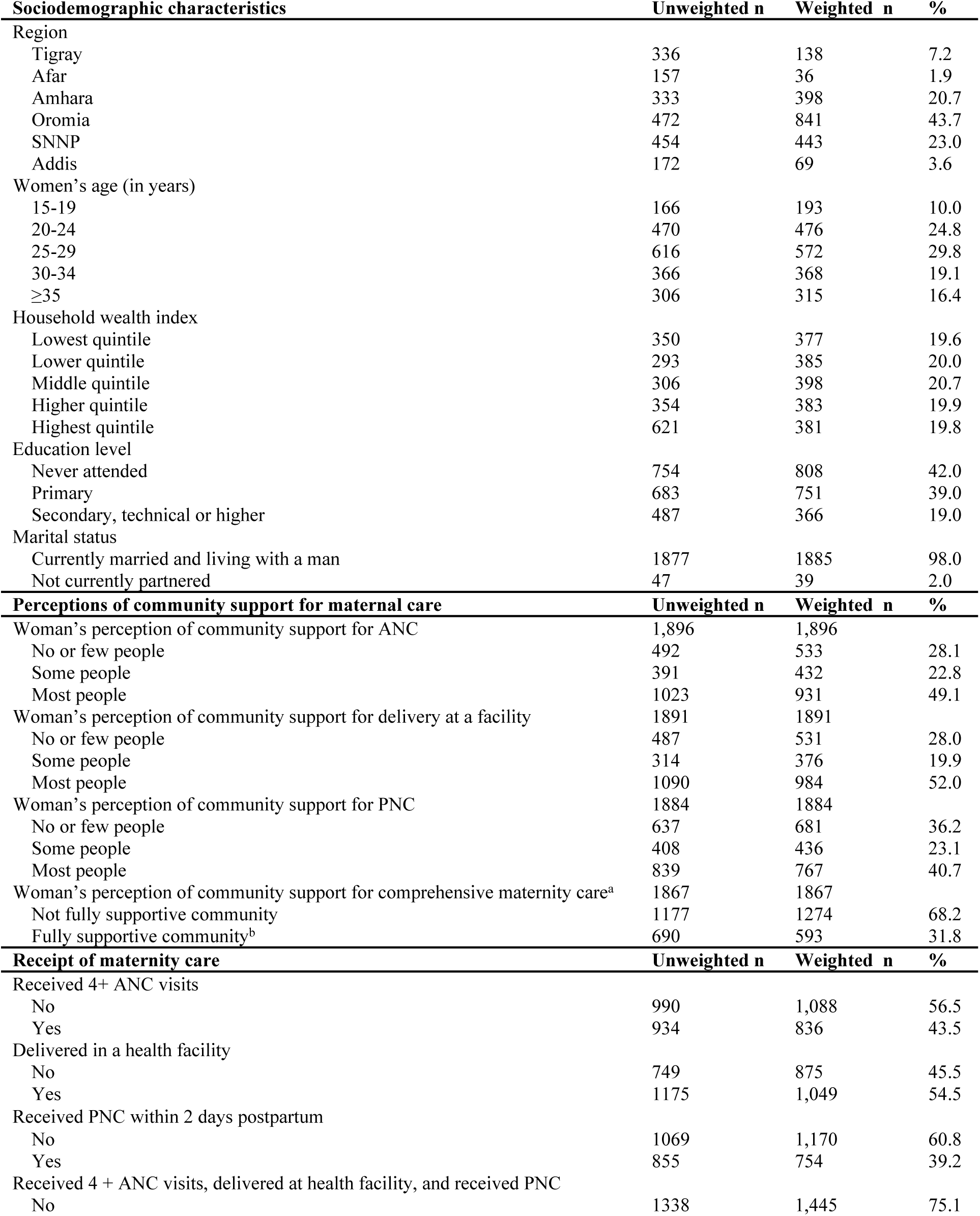
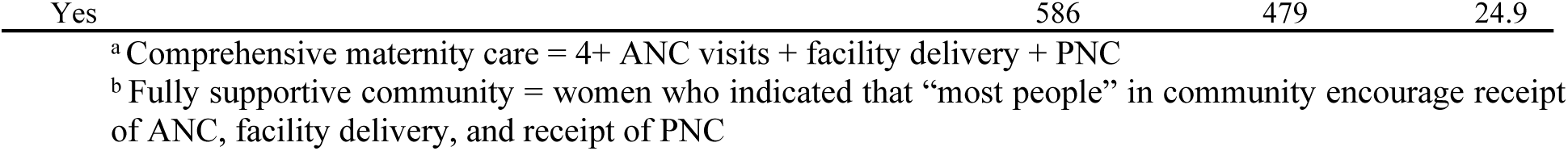
Sociodemographic characteristics of respondents (n=1,924)

Results from the logistic regression analysis examining the association of community support for maternity services with women’s utilization of such services are presented in Table 2. Across all components of the continuum of care, greater perceived support was associated with higher odds of completing each component. For example, women who felt that some people in the community encouraged the use of ANC had 2.35 times higher odds (95% CI: 1.70-3.26) of completing 4+ ANC visits and women who felt that most people encouraged utilization of ANC had more than three times higher odds compared to women who felt there was less community support for ANC (aOR: 3.15, 95% CI: 2.37-4.17). These findings were similar for both facility delivery and postnatal care, with progressively higher odds associated with more perceived support. Lastly, women living in communities that they perceived to be “fully supportive” of encouraging comprehensive maternity care were about twice as likely to receive comprehensive maternity care than women who perceived that the community was not fully supportive of perinatal care (aOR: 1.89, 95% CI: 1.49-2.38).

**Table 2.**
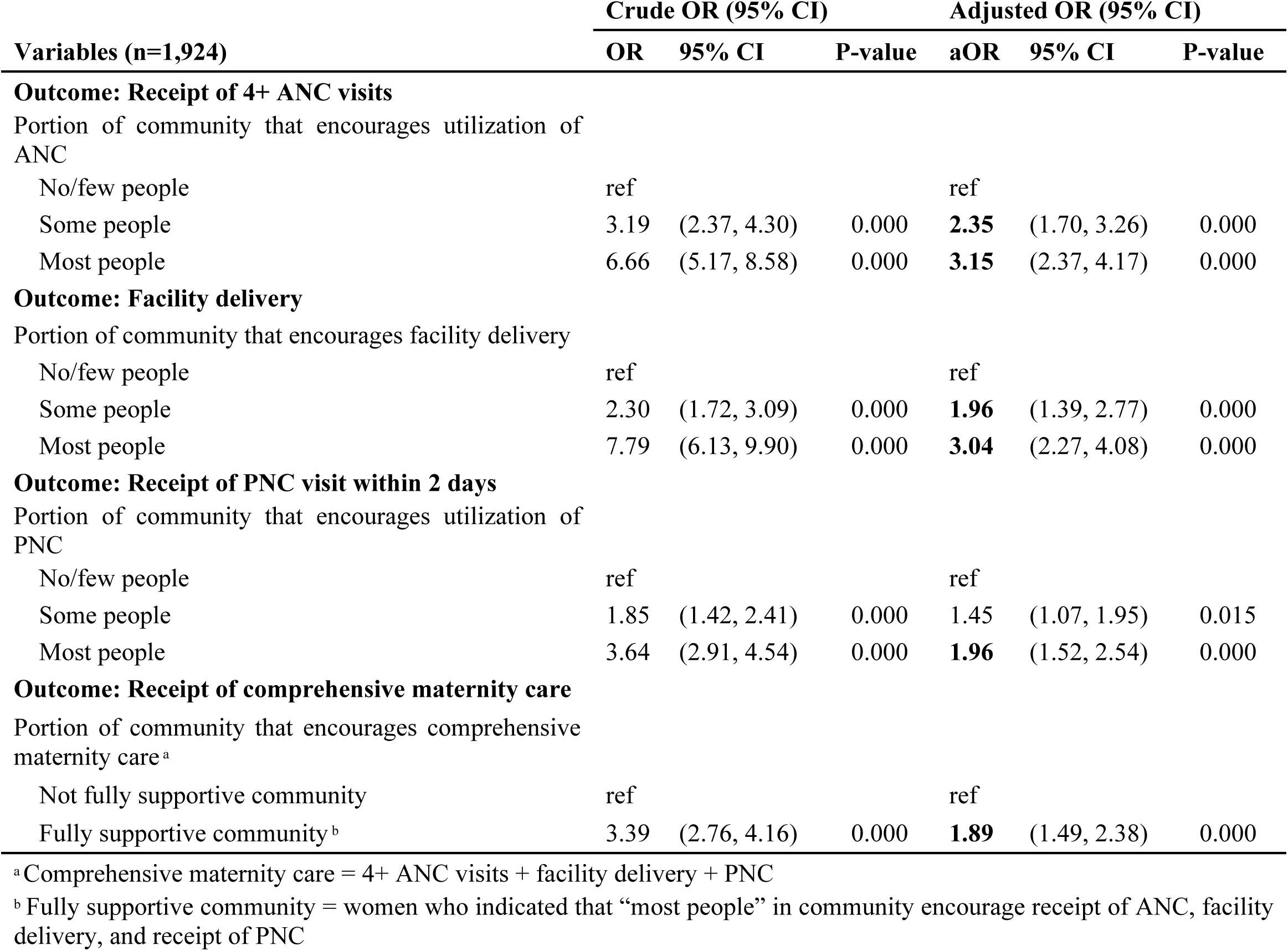
Association between perceived community support for maternity health services with women’s utilization of such services.

| Variables (n=1,924) | Crude OR (95% CI) |  |  | Adjusted OR (95% CI) |  |  |
| --- | --- | --- | --- | --- | --- | --- |
|  | OR | 95% CI | P-value | aOR | 95% CI | P-value |
| **Outcome: Receipt of 4+ ANC visits** |  |  |  |  |  |  |
| Portion of community that encourages utilization of ANC |  |  |  |  |  |  |
| No/few people | ref |  |  | ref |  |  |
| Some people | 3.19 | (2.37, 4.30) | 0.000 | **2.35** | (1.70, 3.26) | 0.000 |
| Most people | 6.66 | (5.17, 8.58) | 0.000 | **3.15** | (2.37, 4.17) | 0.000 |
| **Outcome: Facility delivery** |  |  |  |  |  |  |
| Portion of community that encourages facility delivery |  |  |  |  |  |  |
| No/few people | ref |  |  | ref |  |  |
| Some people | 2.30 | (1.72, 3.09) | 0.000 | **1.96** | (1.39, 2.77) | 0.000 |
| Most people | 7.79 | (6.13, 9.90) | 0.000 | **3.04** | (2.27, 4.08) | 0.000 |
| **Outcome: Receipt of PNC visit within 2 days** |  |  |  |  |  |  |
| Portion of community that encourages utilization of PNC |  |  |  |  |  |  |
| No/few people | ref |  |  | ref |  |  |
| Some people | 1.85 | (1.42, 2.41) | 0.000 | 1.45 | (1.07, 1.95) | 0.015 |
| Most people | 3.64 | (2.91, 4.54) | 0.000 | **1.96** | (1.52, 2.54) | 0.000 |
| **Outcome: Receipt of comprehensive maternity care** |  |  |  |  |  |  |
| Portion of community that encourages comprehensive maternity care<sup>a</sup> |  |  |  |  |  |  |
| Not fully supportive community | ref |  |  | ref |  |  |
| Fully supportive community<sup>b</sup> | 3.39 | (2.76, 4.16) | 0.000 | **1.89** | (1.49, 2.38) | 0.000 |

<sup>a</sup> Comprehensive maternity care = 4+ ANC visits + facility delivery + PNC
<sup>b</sup> Fully supportive community = women who indicated that "most people" in community encourage receipt of ANC, facility delivery, and receipt of PNC

Table 3 demonstrates if and how these effects differ based on urban and rural residence. For ANC, there are significant differences in utilization based on perceived support (aOR_some_:3.84, 95%CI: 1.98-7.43 and aOR_most_: 6.18, 95% CI: 3.55-10.78) and between urban and rural women (aOR_rural_:0.42: 95% CI: (0.24-0.75), however, there is no evidence that the effect of perceived support differs by residence based on the interaction terms (aOR_some*rural_ 0.67, 95% CI: 0.32-1.42 and aOR_most*rural_ 0.68, 95% CI: 0.36-1.27). There is a similar relationship for facility delivery, though the statistical significance among women who perceive some support is attenuated.

**Table 3.** Interaction effects of community perceptions and residence on maternity care utilization.

| Variable | OR | 95% CI | P-value |
| --- | --- | --- | --- |
| **Outcome: Receipt of 4+ ANC visits** |  |  |  |
| Portion of community that encourages utilization of ANC |  |  |  |
| No/few people | reference |  |  |
| Some people | 3.84 | (1.98, 7.43) | 0.000 |
| Most people | 6.18 | (3.55, 10.78) | 0.000 |
| Residence |  |  |  |
| Urban | reference |  |  |
| Rural | 0.42 | (0.24, 0.75) | 0.003 |
| Interaction term |  |  |  |
| Some people * rural | 0.67 | (0.32, 1.42) | 0.298 |
| Most people * rural | 0.68 | (0.36, 1.27) | 0.227 |
| **Outcome: Receipt facility delivery** |  |  |  |
| Portion of community that encourages utilization of facility delivery |  |  |  |
| No/few people | reference |  |  |
| Some people | 3.23 | (0.97, 10.74) | 0.056 |
| Most people | 5.54 | (2.53, 12.12) | 0.000 |
| Residence |  |  |  |
| Urban | reference |  |  |
| Rural | 0.06 | (0.03, 0.13) | 0.000 |
| Interaction term |  |  |  |
| Some people * rural | 0.67 | (0.19, 2.34) | 0.529 |
| Most people * rural | 0.80 | (0.35, 1.83) | 0.593 |
| **Outcome: Receipt PNC within 2 days** |  |  |  |
| Portion of community that encourages utilization of PNC within 2 days of delivery |  |  |  |
| No/few people | reference |  |  |
| Some people | 0.83 | (0.49, 1.40) | 0.477 |
| Most people | 1.37 | (0.89, 2.11) | 0.150 |
| Residence |  |  |  |
| Urban | reference |  |  |
| Rural | 0.10 | (0.06, 0.15) | 0.000 |
| Interaction term |  |  |  |
| Some people * rural | 2.60 | (1.39, 4.84) | 0.003 |
| Most people * rural | 2.27 | (1.34, 3.85) | 0.002 |
| **Outcome: Receipt of comprehensive maternity care** |  |  |  |
| Portion of community that encourages comprehensive maternity care* |  |  |  |
| Not fully supportive community | reference |  |  |
| Fully supportive community** | 1.81 | (1.33, 2.45) | 0.000 |
| Residence |  |  |  |
| urban | reference |  |  |
| rural | 0.15 | (0.132, 2.45) | 0.000 |
| Interaction term |  |  |  |
| Some people * rural | reference |  |  |
| Most people * rural | 1.57 | (1.00, 2.45) | 0.049 |

There is evidence of interaction between community support and residence for both receipt of PNC and the full continuum of care, however. When accounting for interaction, the main effect of community support on PNC utilization is no longer significant - among urban women, there is no difference in the odds of receiving PNC by level of perceived community support. The main effect of residence remains significant; among women who perceive little support in the community, those who live in rural areas have significantly lower odds of utilizing PNC than women who live in urban areas (aOR:0.10, 95% CI: 0.06-0.15). The interaction, however, demonstrates that the effect of community support is significantly stronger among rural women, with perceived levels of community support being strongly associated with increased odds of PNC utilization (aOR_some*rural_ 2.60, 95% CI: 1.39-4.84 and aOR_most*rural_ 2.27, 95% CI: 1.34-3.85). Regarding the full continuum of care, the main effects of both perceived support (aOR_full support_ 1.81, 95% CI: 1.33-2.45) and residence (aOR_rural_ 0.15, 95% CI: 0.13-2.45) retain statistical significance; among urban women, women who perceive full support are significantly more likely to receive all components of care while among women who perceive low support, rural women are significantly less likely to receive all components relative to urban women. As with PNC utilization, the effect of perceiving full support is stronger among rural women than among urban women (aOR_full support*rural_ 1.57, 95%CI: 1.00-2.45).

## Discussion

To our knowledge, this is the first study to quantitatively examine the association between women’s perception of community support for and utilization of maternity healthcare services across the continuum of care in Ethiopia. We find that perceived support from the community significantly influences the likelihood of receiving care across each component of the continuum and across the full continuum, with the effect being stronger amongst rural women for completion of PNC and the full continuum.

Our findings suggest that when women feel that at least some people in their community are supportive of maternal health services, they are more likely to receive these services and that this applies across the entire continuum of care. This is consistent with previous qualitative studies indicating that communities have considerable influence on the receipt of services. For example, a study in Addis Ababa highlighted that community members felt there was little utility to receiving ANC services and discouraged women from accessing this care (21). Similarly, women who received poor quality delivery services discouraged others from attending services, a finding which was echoed in a similar study in Kenya (34). These studies, which focus on only one component of the continuum, demonstrate the critical role that community support plays in women’s utilization of specific maternity care services. No studies that we could find examined the influence of community support across all components of the continuum of care, limiting our ability to compare this finding; nonetheless, that each component was consistently associated with higher uptake of services points to the importance of building community support for each service in order to increase utilization of the entire continuum. We also found that, while women generally considered that at least some people in their community encouraged care seeking, between one in four and one in three, depending on the service, reported that either no or few people supported them. This points to the continued need to address community opinions on maternal health services and improve perceptions of the need for and quality of services. Understanding the specific beliefs associated with each service and barriers that prevent engagement is crucial for improving women’s utilization of services.

Additionally, we found that across the entire continuum of services, urban women were consistently more likely to utilize services than rural women, but that the effect of perceived community support was significantly stronger among rural women for receipt of PNC services and for completing the entire continuum. Disparities in use of maternal health services between urban and rural women in Ethiopia have been well documented in other studies (35–37). There are a number of reasons why these disparities exist, including access-related challenges (38,39) and socioeconomic differences such as education and wealth (40,41), but few studies have examined whether and how specific factors, and particularly community support, differ in their effect based on residence. A study in Morocco demonstrated that restrictive gender norms impacted sexual and reproductive healthcare-seeking behavior more strongly for rural than urban women (42), while in Ethiopia, community-level gender norms were significantly related to experience of childhood violence in rural, but not urban, areas (43). While there are no studies that we could find that include a comparative perspective of the influence of community support on maternal health care-seeking by residence, our results indicate that difference do exist and are important to consider, at least for services that are less commonly used such as PNC. Perceptions of community support for postnatal care may be particularly impactful in rural areas, where women traditionally have less autonomy to make individual decisions due to less access to economic resources, and persistence of conservative patriarchal norms, prioritizing husband and extended family decision-making (44). Conversely, in urban areas, women may have greater access to individual financial resources and traditional norms are frequently weaker (43,45), thus the effect of perceived community support may have less influence on individual decisions around care. Understanding community perceptions of the need for PNC care, particularly in rural areas, in order to design effective community-based interventions may serve as a means to reduce disparities in urban and rural utilization.

Our study has several strengths. First, this study used longitudinal data collected amongst a representative sample of currently pregnant women who were asked to report on their perceptions of community support prior to utilization of services (with the potential exception of ANC), which reduces concerns about temporality. Additionally, we utilized a novel measure of perceived community support, asking women to report on the community in general, rather than rely on aggregate measures of individual data, which are generally used to measure community influences (46–48). As these data are generally collected only among women age 15-49, they fail to capture the opinions of other individuals in the community (46), who are likely impactful in determining care-seeking behavior. Finally, we explore the modifying effect of residence on the effect of perceived support, providing more evidence into how mechanisms that influence care seeking may differ between urban and rural women and explain disparities in care. We do note limitations in our data, including that our measure was non-specific in is wording. “Support” can entail any number of actions or behaviors, but we were intentionally vague to allow the respondent to interpret support as she saw fit. Additional research is needed to understand the components of community support and its specific impact on women’s utilization of maternal health services. Additionally, while we collection information from women on their perceptions of community support, we did not collect information from male partners and other community members who influence decisions. Continued research to understand beliefs amongst a more diverse population than women of reproductive age would help to identify barriers to care.

## Conclusions

Perceived community support is an important predictor of women’s utilization of maternal care in Ethiopia. These findings shed light on an important mechanism to care-seeking, and how it differs by urban and rural residence, contributing to ongoing disparities in care between urban and rural women. We note that while support is generally high across the continuum, at least one in four women report that few or no people support care-seeking, highlighting a lack of awareness of services that are important to address. Additional research is needed to better understand the reasons for this lack of support, particularly towards services in the postpartum period when the majority of maternal and newborn mortality occur.

## Data Availability

All data are fully available without restriction

https://www.pmadata.org/data/available-datasets

## Declarations

### Abbreviations

ANC: Antenatal Care
PNC: Postnatal Care
AAU/CHS: Addis Ababa University, College of Health Sciences
JHSPH: Johns Hopkins University Bloomberg School of Public Health
PMA: Performance Monitoring for Action
FMoH: Ethiopian Federal Ministry of Health
EAs: Enumeration Areas

## Ethical declarations

### Ethics approval and consent to participate

PMA Ethiopia received ethical approval from Addis Ababa University, College of Health Sciences (AAU/CHS) (Ref: AAUMF 01-008) and the Johns Hopkins University Bloomberg School of Public Health (JHSPH) Institutional Review Board (FWA00000287). Verbal informed consent was obtained from all of the participants. The IRB approves verbal consent procedures (without a need for written consent) for simple surveys without any invasive procedures in an environment where literacy is low. Women under the age of 18 who are married are considered emancipated minors and are able to provide informed consent. No unemancipated minors were included in this survey. Detailed information about the ethical guidelines can be found in the National Research Ethics Review Guidelines (http://www.ccghr.ca/wp-content/uploads/2013/).

### Consent for publication

Not applicable.

## Availability of data and materials

The datasets generated during the study are publicly available from the PMA website (https://www.pmadata.org/data/available-datasets).

## Acknowledgments

The PMA Ethiopia project relies on the work of many individuals, both in the United States and in survey countries. Thanks to the Ethiopia country team and resident enumerators who are ultimately responsible for the success of PMA Ethiopia.

## Footnotes

^1^Initial study design and sampling took place prior to the creation of three new regions from within SNNPR and thus represents estimates within the previous administrative borders of SNNP.

